# Developing and deploying a use-inspired metapopulation modeling framework for detailed tracking of stratified health outcomes

**DOI:** 10.1101/2025.05.05.25327021

**Authors:** Arindam Fadikar, Abby Stevens, Sara Rimer, Ignacio Martinez-Moyano, Nicholson Collier, Jonathan Ozik, Charles Macal

## Abstract

Public health experts studying infectious disease spread often seek granular insights into population health outcomes. Metapopulation models offer an effective framework for analyzing disease transmission through subpopulation mixing. These models strike a balance between traditional, homogeneous mixing compartmental models and granular but computationally intensive agent-based models. In collaboration with the Chicago Department of Public Health (CDPH), we developed `MetaRVM`, an open-source R package for modeling the spread of infectious diseases in subpopulations, which can be flexibly defined by geography, demographics, or other stratifications. `MetaRVM` is designed to support real-time public health decision-making and through its co-creation with CDPH, we have ensured that it is responsive to real-world needs. We demonstrate its flexible capabilities by tracking influenza dynamics within different age groups in Chicago, by integrating an efficient Bayesian optimization-based calibration approach.

## 1 INTRODUCTION

In the wake of COVID-19, public health decision-makers have come to recognize epidemiological modeling as an indispensable part of a robust response strategy [21]. Although traditional surveillance dashboards can reveal in real time how an outbreak is unfolding, epidemiological models reach further by forecasting likely futures, stress-testing policy interventions, and guiding the strategic allocation of limited resources. In Chicago, this recognition catalyzed a formal collaboration between the Chicago Department of Public Health (CDPH), computational scientists at Argonne National Laboratory, and epidemiology researchers at the University of Chicago to embed epidemiological modeling directly into the department’s routine surveillance workflow. Our partnership follows a *co-creation* approach to ensure that the analytical tools we design are scientifically sound and also immediately operational [15].

Epidemiological modeling sits on a spectrum. At one end are classic Susceptible–Infected–Recovered (SIR) compartmental models, which are systems of ordinary differential equations that assume a single, well-mixed population and require minimal computation. At the other are agent-based models (ABMs), which track every person in a synthetic population and explicitly simulate their contact patterns, behaviors, and environments, enabling highly granular insights but requiring significant compute power. The urgency of public health needs during the COVID-19 pandemic and the unprecedented data availability that accompanied it spurred the development of many advanced epidemiological modeling frameworks [10, 27], many of which sought to strike a balance between computational complexity and modeling granularity. Prominent examples include `flepiMoP` [11], an open-source workflow that automates model specification and calibration for state- and county-level decision makers, and mobility-driven metapopulation systems that embed commuting flows in transmission dynamics [2, 19].

Our team previously built CityCOVID [17, 13], a city-scale ABM built on the ChiSIM framework [12] that represents every Chicago resident as an individual software agent following a realistic daily schedule. CityCOVID generated high-resolution projections and quantified the effects of non-pharmaceutical interventions, such as mask mandates and school closures, for the Chicago and Illinois Departments of Public Health [8]. While these outputs were richly granular and policy-relevant, they came at a price: CityCOVID demands high-performance computing and expert staff, making it impractical for routine use within CDPH’s analytics pipeline. On the other hand, an off-the-shelf SIR model, though fast and easy to deploy, ignores critical heterogeneity (such as age structure, neighborhood context, and heterogeneous contact patterns) that shapes both exposure risk and clinical severity in a city as large and diverse as Chicago.

In an effort to minimize computational complexity while still leveraging the rich granularity of CityCOVID, we developed `MetaRVM` [3], an open source `R` package designed to support quick turnaround public health decision-making for infectious disease outbreaks. `MetaRVM` is a *metapopulation model* [25, 26], *a type of model that occupies a middle ground between simple compartmental models and ABMs. They extend the classic SIR framework by propagating infection across interacting subpopulations* (e.g., age groups, neighborhoods, occupations), whose interactions are governed by realistic contact networks. `MetaRVM` allows users to model subpopulations defined along any strata of interest, and thanks to the rich synthetic population our team has developed and maintained as part of CityCOVID, we are able to generate contact networks between many different possible subpopulations in the city of Chicago.

`MetaRVM` was developed in close collaboration with CDPH, and its design and implementation are oriented toward usability, flexibility (in the diseases it supports and the subpopulations of interest), and interpretability of results and insights. In addition to releasing `MetaRVM` as an open-source `R` package, we have also developed a front-end `shiny` interface that allows users to easily upload model parameters and specifications, run the model, and generate a variety of figures to quickly assess results. We demonstrate the core functionalities of `MetaRVM` through the example of tracking influenza-related hospitalizations in Chicago for the 2023-24 flu season, a use-case that highlights both the challenges and opportunities of incorporating epidemiological modeling into public health workflows.

## 2 MetaRVM

This section provides a comprehensive description of the `MetaRVM` modeling framework, which is implemented as an open source R package with a front-end `shiny` interface. As shown in Figure 1, at each simulation step, the population is divided into user-defined subpopulations that interact through time-varying mixing matrices that are derived from a synthetic population. Infections progress through each subpopulation using an extended Susceptible-Exposed-Infected-Recovered (SEIR) framework that accounts for vaccination, detailed disease states, and mixing between subpopulations. Here, we present the structure of the model, the compartmental transitions, and the key mathematical formulations underlying the simulation engine.

**Figure 1:**
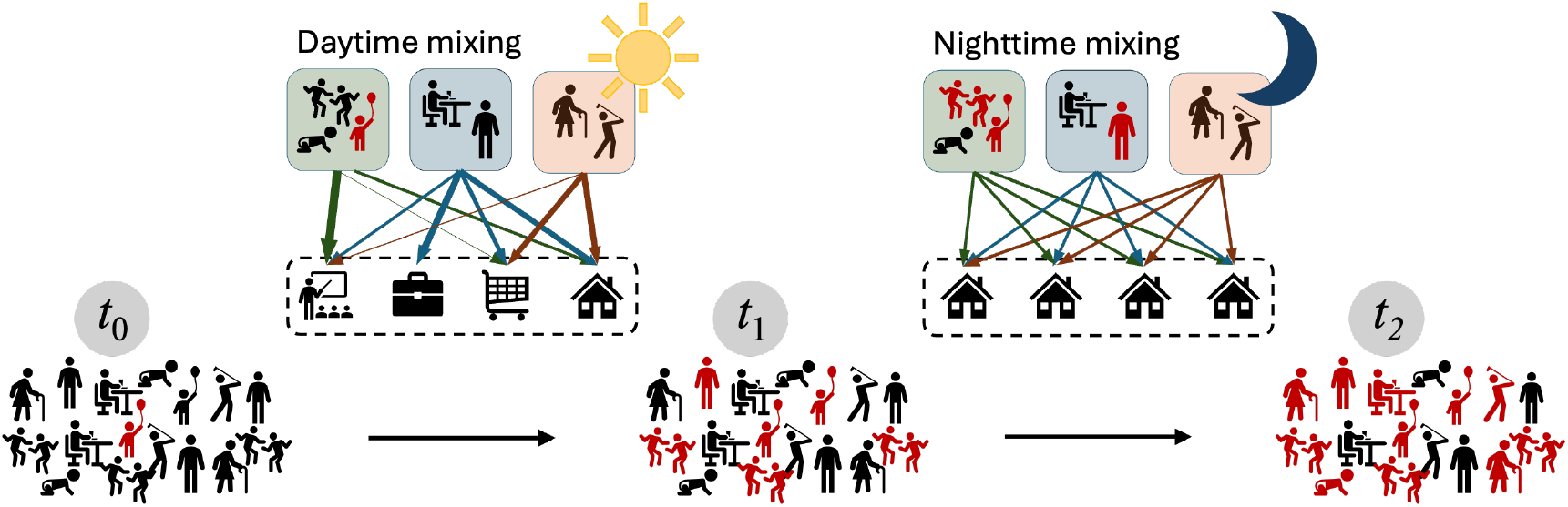
Conceptual overview of the `MetaRVM` metapopulation engine, with exemplar subpopulations based on age: children (green boxes), working-age adults (blue boxes), and seniors (orange boxes). During the daytime, these subpopulations mix across multiple locations (schools, workplaces, stores, households), with colored arrows indicating the frequency of visits for each subpopulation. During the nighttime, mixing takes place almost exclusively within households. Infection can propagate whenever susceptible (black) and infectious (red) individuals are in contact with one another. The population is initialized with a single infection at time *t*_0_, which then spreads to student and teachers during the school day by time *t*_1_, and finally to other subpopulations overnight by time *t*_2_.

**Figure 2:**
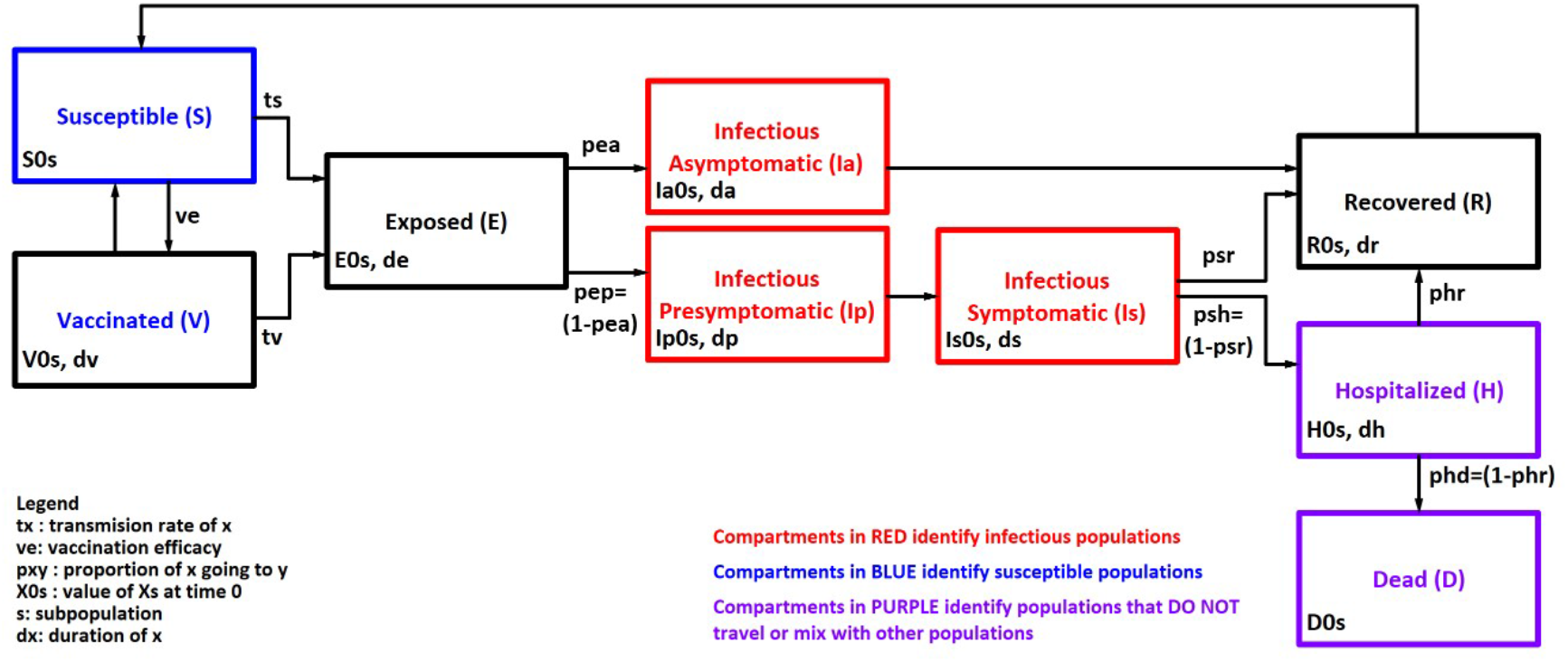
The state chart of the MetaRVM model

### 2.1 Model Structure

The `MetaRVM` model builds upon the SEIR framework by introducing additional compartments to capture more detailed dynamics of disease progression, while allowing for heterogeneous mixing among different demographic stratum. These generalizations allow the model to account for factors such as vaccinations, hospitalizations, and fatalities. At the start of a simulation, nearly all individuals in the population are classified as Susceptible (S), except for the initial case of infection, often referred to as “patient zero”. If a portion of the population is vaccinated, those individuals are moved to the Vaccinated (V) compartment. The degree of immunity conferred by vaccination depends on the vaccine efficacy parameter (*ve*). When susceptible individuals come into contact with infectious individuals, disease transmission occurs based on the probability of transmission. The calculation of effective transmission probabilities in multi-population scenarios is detailed in the next subsection. Importantly, vaccinated individuals can still be exposed to the disease, albeit with a reduced probability, especially if the vaccine does not provide complete immunity. Additionally, immunity from vaccination wanes over time at a rate of 1*/dv*, where *dv* represents the average duration of vaccine-conferred immunity (in days). Once exposed to the infection, the susceptible and vaccinated individuals transition to the Exposed (E) state. The incubation period, after which exposed individuals become infectious, lasts an average of *de* days. `MetaRVM` model distinguishes between asymptomatic and symptomatic infectious states, as these groups differ in disease progression and their interactions with the broader population. After the incubation period, exposed individuals move to the infectious state, with the proportion entering the asymptomatic infectious state (Ia) determined by the parameter *pea*. The remainder transition to the presymptomatic infectious state (Ip). Individuals remain in these states for an average of *da* and *dp* days, respectively. Asymptomatic individuals recover after their infectious period, while presymptomatic cases progress to the symptomatic infectious state (Is). From there, individuals either recover (R) or require hospitalization (H), with transitions occurring at rates of 1*/dp* and 1*/ds*, respectively. The fraction of symptomatic individuals who recover directly is governed by the parameter *psr*, while the remainder are hospitalized. Hospitalized individuals stay in the H compartment for an average duration controlled by the parameter *dh*. From the hospital, individuals either recover or succumb to the disease (D), with the proportion of fatalities determined by the parameter *phd*. For diseases where reinfection is possible, individuals in the Recovered (R) compartment may return to the Susceptible (S) state after an average of *dr* days. This structure of the disease progression in `MetaRVM` allows it to adapt to many common respiratory diseases.

### 2.2 Probability of Exposure

The probabilities of a susceptible or vaccinated individual transitioning to the exposed state are defined as follows:

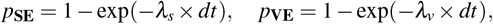

where *λ_s_* and *λ_v_* represent the force of infection for susceptible and vaccinated individuals, respectively, and *dt* is the discrete time step in the simulation. The force of infection depends on the contact rates (commonly denoted as *β* ), vary across subpopulations and are calculated based on mixing matrices, which account for interactions between different population strata. Specifically, infected individuals in stratum *j* can mix with individuals in stratum *j^′^*, and vice versa, modifying the force of infection within each subpopulation. However, individuals in the Hospitalized (H) and Deceased (D) states are excluded from the effective population that interacts with other subpopulations.The total effective population, denoted as **MP**, excludes individuals in the H and D states. It is calculated as:

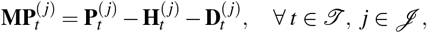

where 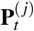 represents the total population in stratum *j* at time *t*, and 𝒥 is the set of all demographic strata. Next, let *M* denote the mixing matrix of order 𝒥 × 𝒥 , where the (*i, j*)th element *m_i j_* represents the fraction of the population in stratum *i* that mixes with individuals in stratum *j*. The matrix satisfies the condition:

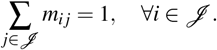

Using the mixing matrix, we calculate the effective population size and the effective number of infectious individuals for each stratum at any given time *t* as:

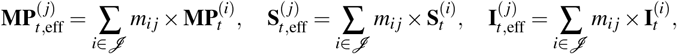

where 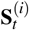 and 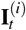 represent the susceptible and infectious populations in stratum *i* at time *t*, respectively. Using these effective values, the force of infection for susceptible and vaccinated individuals in each stratum is calculated as:

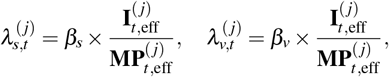

where *β_s_* and *β_v_* are transmission scaling factors for susceptible and vaccinated individuals, respectively. To determine the number of transitions from the Susceptible (S) and Vaccinated (V) compartments to the Exposed (E) compartment, the stratum-specific force of infection is applied. First, the distribution of the susceptible population across strata is calculated using the mixing matrix. Then for each stratum *i*, the number of susceptible individuals transitioning to the exposed state is given by:

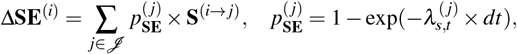

where **S**^(*i*→*j*)^ represents the portion of the susceptible population in stratum *i* that mixes with stratum *j*, and 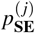 is the probability of exposure in stratum *j*. Similarly, the number of vaccinated individuals transitioning to the exposed state is calculated as:

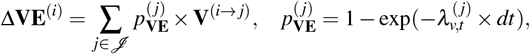

where **V**^(*i*→*j*)^ represents the vaccinated population in stratum *i* that mixes with stratum *j*, and 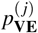 is the probability of exposure in stratum *j*.

### 2.3 Generating mixing matrices with the ChiSIM synthetic population

Mixing matrices translate where and how often people meet into quantitative terms the model can use: each cell *m_i j_* records the expected number of potentially infectious contacts that a member of stratum *i* has with members of stratum *j* per unit time. Because the force of infection is proportional to these contacts, estimating them accurately is highly consequential to any metapopulation model. Mixing matrices can be derived analytically using gravity or radiation models, which require minimal input such as population counts and geographic centroids [1, 23]. However, as rich empirical data streams become increasingly available, it has become common to derive contact patterns directly from data [7]. Social contact surveys such as POLYMOD [14], commuting flows from the American Community Survey (ACS) [24], and even cell phone mobility data [28, 26] have all been used to generate empirical mixing matrices.

#### Synthetic population

Here, we propose an alternative that leverages the rich synthetic ecosystem our group has been developing and maintaining for many years. Underlying CityCOVID, and the ChiSIM ABM more broadly, is a synthetic population [9] that is statistically representative of Chicago’s population (2.7 million persons), along with their associated places (1.4 million locations) and behaviors (13,000 activity schedules). During a simulated day, agents move from place-to-place, hour-by-hour, engaging in social activities and interactions with other colocated agents, resulting in an endogenous co-location or contact network. We use data from ACS and the Public Use Microdata Sample (PUMS) to generate geolocated households of individuals that are statistically representative of both individual and household demographics at the census block group (CBG) level. Workplaces are synthetically generated to match Census County Business Patterns data at the ZIP code level, and individuals identified as employed are assigned a workplace based on the LEHD Origin-Destination Employment Statistics at the CBG level. Schoolaged children are assigned an actual school in Chicago, and a subset of working adults have schools assigned as their workplaces. In addition, our synthetic ecoystem includes a variety of other locations where social mixing occurs, such as restaurants and gyms, which were derived from the SafeGraph dataset. Individuals move between locations according to assigned hourly schedules from the American Time Use Survey (for adults) and the Panel Study of Income Dynamics (for children) based on their matching demographic characteristics. To capture the inherent variability of human behavior and mobility patterns, individuals are assigned ten different schedules for both weekdays and weekends, and each day in the simulation randomly selects one from the appropriate category.

#### Counting contacts

Let *N* denote the number of individuals in the population, **L** the set of possible locations, and **T** the time periods over which we wish to estimate mixing patterns. We partition the population into subpopulations 𝒥 (e.g., age groups, neighorbhoods, income brackets) and let *n* _j_ represent the number of individuals in subpopulation *j*, where ∑ _*j* ∈ 𝒥_*n _j_* = *N*. To generate mixing matrices using the synthetic population, we use the following contact counting method. Suppose an individual belongs to subpopulation *j* ∈ 𝒥 . We randomly select an available activity schedule for this individual, which tells us they will be at location *l* ∈ **L** at time *t* ∈ { 1*, … , T* }. Repeating this for all individuals in the population, we are able to count the total number of people from each subpopulation who were *co-located* at location *l* at time *t*, which we denote *n _j_*(*l, t*) for *j* ∈ 𝒥 .

A *contact* between individuals occurs when they are at the same place at the same time. After assigning all individuals a location at time *t*, we count the total number of contacts that occur at each location *between* and *within* subpopulations. Between subpopulations, the total number of contacts is counted as the number of possible pairs between the *n _j_*(*l, t*) people in *j* and the *n_i_*(*l, t*) people in *i*, given by *C_i j_*(*l, t*) = *n_i_*(*l, t*)*n_j_*(*l, t*). Within subpopulation *j*, we exclude self-contacts and have *C_j j_*(*l, t*) = (*n_j_*(*l, t*))(*n_j_*(*l, t*) − 1). The mechanics of our population and location assignments ensure that individuals are separable across space and time, hence we can estimate the overall proportion of contacts between subpopulations *j* and *i* relative to the total number of contacts incurred by subpopulation *j*, or entry *m_i j_* in mixing matrix *M*, by

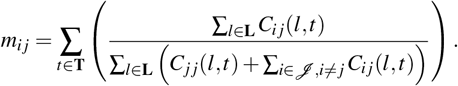

### 2.4 MetaRVM R Package

The `MetaRVM` [3] model is implemented as an open-source package in the R programming language [20], leveraging the `odin` library [6] for model specification and simulation. `Odin` is a domain-specific language (DSL) embedded within R that facilitates the definition of compartmental models as systems of ordinary differential equations (ODEs). One of the key advantages of `odin` is its ability to translate high-level ODE specifications into optimized C code, which is then compiled and executed from within the R environment. This compilation significantly accelerates model simulations, making it suitable for applications that require repeated model evaluations, such as calibration, uncertainty quantification, and scenario projections.

The core simulation engine of `MetaRVM` supports both deterministic and stochastic simulation modes. To promote usability and reproducibility, `MetaRVM` allows users to define model parameters and initial conditions through a structured YAML configuration file. This parameter file can be supplied either via command-line execution or through an interactive interface provided by a `shiny` dashboard, which is bundled with the package. The dashboard enables non-programmers, including public health practitioners and policy analysts, to run simulations, visualize outputs, and explore different scenarios without writing code.

## 3 Tracking Influenza Hospitalizations in Chicago

To demonstrate the use of `MetaRVM`, we apply the model to prospective surveillance of influenza-related hospitalizations in the city of Chicago during the 2023–24 influenza season, commencing in October. This case study is motivated by the operational needs of public health agencies, which aim to anticipate hospital burden, characterize transmission dynamics across demographic strata, and allocate healthcare resources in a temporally responsive manner. This task is inherently challenging due to data sparsity, reporting lags, and the difficulty of accurately modeling epidemic trajectories at the subpopulation level. Leveraging the data available for the 2023–24 season, we configured `MetaRVM` to jointly model hospitalization incidence across three age-stratified groups: 0–17, 18–26, and 65 years and older.

### 3.1 Data

#### Influenza hospitalizations

The city of Chicago does not publish data on reported influenza hospitalizations, but it does report the weekly number of influenza-associated ICU hospitalizations [16]. We estimate the total number of hospitalizations from ICU admissions using age-stratified ratios derived from a recent study of the rates of influenza-associated hospitalizations and ICU admissions from 2009 to 2019 [18], specifically using rates reported in eTable 2. We scaled the weekly reported ICU admissions according to this derived ratio for each age group to use as a proxy for hospitalizations.

#### Mixing matrices

To account for the temporal variation in contact behavior relevant to infectious disease transmission, four age-structured mixing matrices were developed, stratified by three demographic groups: children and adolescents (0–17 years), adults (18–64 years), and older adults (65 years and above). This particular stratification was motivated by the availability of influenza vaccination coverage data, which was reported at these three demographic levels. Aligning the model structure with available immunization data ensures consistency between behavioral assumptions and intervention scenarios, particularly for evaluating age-specific vaccination impacts.

The mixing matrices further distinguish contacts across two temporal dimensions—weekday vs. weekend and daytime vs. nighttime—resulting in four distinct contact settings. As shown in Figure 3a, contact patterns exhibit clear temporal and demographic structure. During weekday daytime periods, individuals primarily interact within their own age group, especially among children and working-age adults. This pattern reflects age-assortative mixing driven by structured environments such as schools and workplaces. In contrast, weekday nighttime periods reveal more heterogeneous mixing across age groups, consistent with household-based interactions that typically involve individuals from multiple age cohorts. On weekends, contact patterns also differ substantially from weekday structures (not shown here). The 0–17 age group, in particular, shows reduced within-group contact during both daytime and nighttime, suggesting lower engagement in structured peer interactions and increased intergenerational contact. These findings highlight how social schedules, institutional settings, and household composition influence age-specific mixing patterns—factors that are critical for accurately modeling pathogen transmission, estimating reproduction numbers, and designing effective public health interventions.

#### Vaccination

We obtained the historical vaccination data for the year 2023-24 from the city of Chicago data portal which reports daily number of vaccinations for these three age-groups. Figure 3b shows the daily vaccination rates in each of the population strata. We noted significantly higher vaccination rates in the older population.

**Figure 3:**
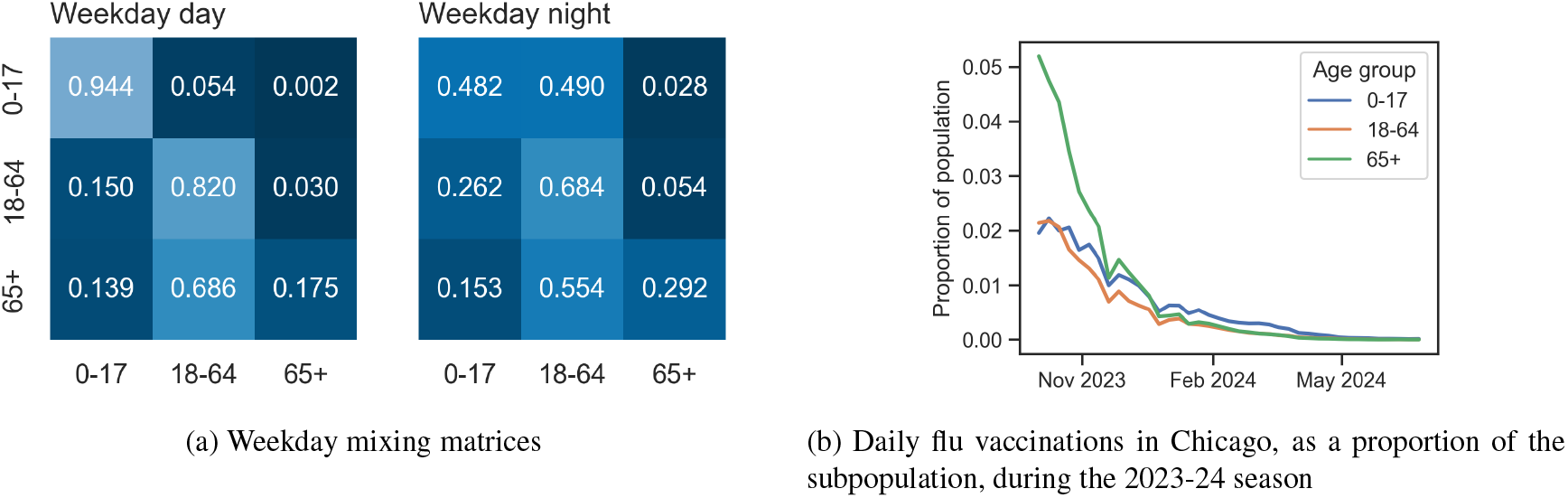
Mixing matrices and vaccination records, stratified by age groups, supplied to `MetaRVM` for modeling influenza-related hospitalizations in Chicago during the 2023-24 flu season.

### 3.2 Calibration

We calibrate the `MetaRVM` model to weekly estimates of influenza-related hospitalizations in three sequential phases. This phased calibration accounts for the time-varying nature of transmission dynamics and public health interventions, enabling a more flexible fit to the data over time [5]. In the first phase, the model is initialized and fitted to the first 10 weeks of estimated hospitalization counts. A parameter search is conducted to generate plausible sets of model parameters, and corresponding epidemic trajectories. The most promising parameter set producing a realistic trajectory is selected, and the model state is checkpointed at the end of week 10. This checkpointing mechanism preserves the internal compartments, and transmission history of the model, allowing consistent resumption of simulations in later phases.

The second phase begins by resuming the simulation from the week-10 checkpoint and extends the calibration window to include weeks 11 through 20. Recognizing that epidemic dynamics are non-stationary due to factors such as behavioral changes, policy interventions, and waning immunity, model parameters are re-estimated during this interval. As in the first phase, the best-fitting trajectory is identified and the model is checkpointed at the end of week 20. The third phase repeats this procedure for subsequent time periods, allowing the model to adapt sequentially to evolving trends.

In each of the calibration phases, we estimate four key epidemiological parameters for each of the three sub-populations considered in the model, resulting in a total of 12 parameters per phase. These parameters include: (1) the transmission rate among susceptible individuals (*β_s_*), (2) the transmission rate among vaccinated individuals (*β_v_*), (3) the proportion of asymptomatic infections among all infectious cases (*pea*), and (4) the proportion of symptomatic cases that require hospitalization (*psh*). These parameters are critical for capturing the heterogeneity in transmission and disease severity across different population strata. Other model parameters were kept at fixed values obtained from literature.

To efficiently explore the high-dimensional parameter space, we use Bayesian Optimization (BO) in conjunction with Thompson Sampling [4]. BO offers a principled, sample-efficient approach to global optimization by constructing a surrogate probabilistic model—typically a Gaussian Process (GP) [22]—that approximates the unknown objective function. In our context, this objective function quantifies the discrepancy between simulated and observed hospitalization counts. The GP surrogate provides predictions of the function values at unsampled points in the parameter space along with the associated uncertainties.

Thompson Sampling is adopted as the acquisition strategy within the BO framework. At each iteration, a sample is drawn from the posterior distribution using the surrogate model, and the parameter configuration that maximizes the sampled function is selected for evaluation. This strategy effectively balances exploration, sampling in regions of high uncertainty, and exploitation, focusing on regions likely to yield better fits, without the need for heuristic tuning of exploration-exploitation trade-offs. The selected parameter configurations are then passed to the full MetaRVM simulator, which evaluates the model by comparing the resulting epidemic trajectory against the observed data. The results of each simulation are fed back to update the surrogate model at each iteration. This closed-loop, active learning approach significantly reduces the number of model evaluations required to find plausible parameter in the high-dimensional space. Figure 4a and Figure 4b present the fitted `MetaRVM` trajectory summaries and the posterior distributions of the transmissibility parameter *β_s_*, respectively. Notably, the distribution of *β_s_* evolves over time as the flu season progresses, reflecting changes in transmission dynamics. Capturing this temporal variation within the modeling and calibration framework is a key component here, as it can provide policymakers with more accurate and adaptive insights for decision-making throughout the epidemic.

**Figure 4:**
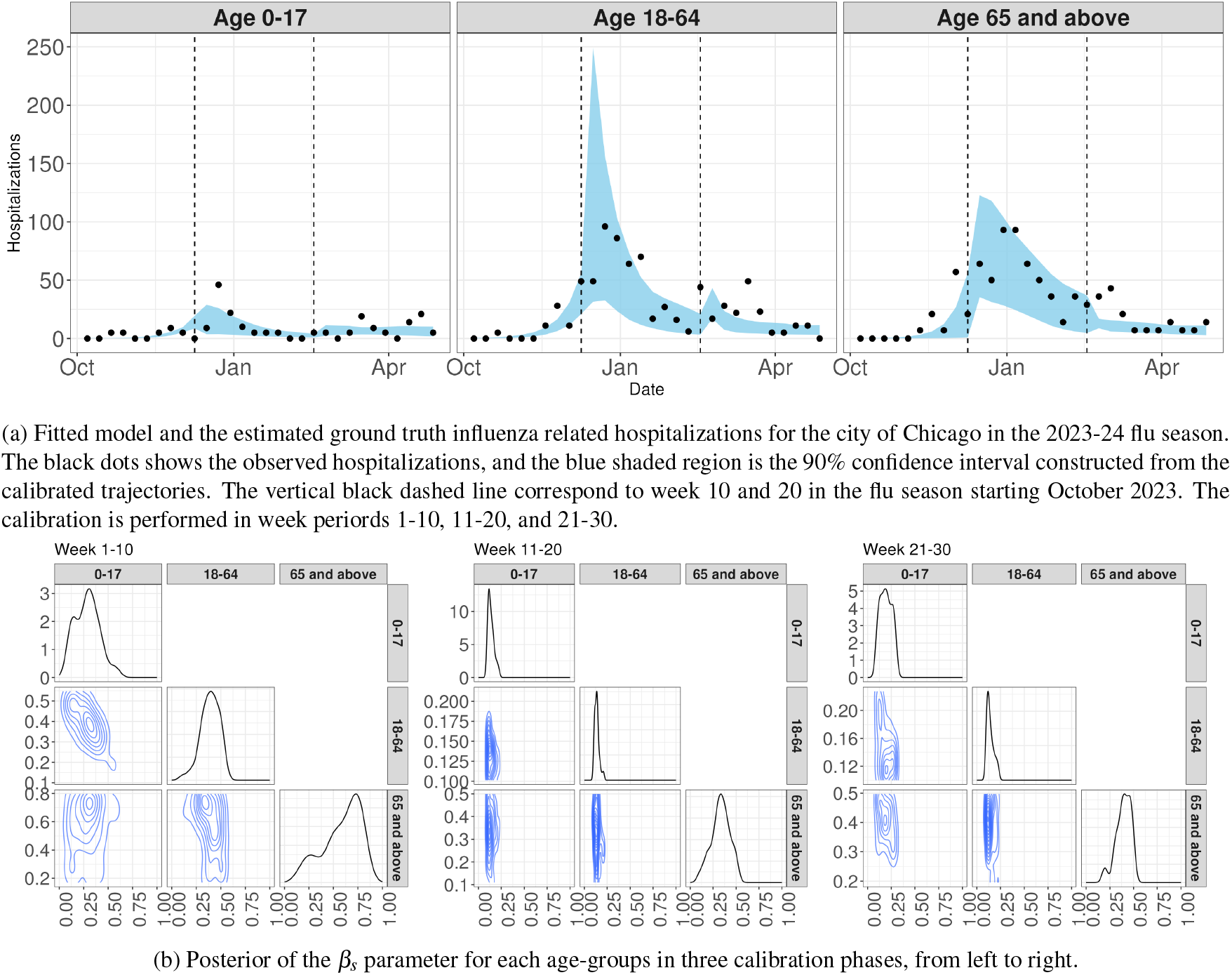
Model calibration to the influenza related hospitalizations in Chicago during 2023-24 flu season.

## Data Availability

All data produced in the present work are contained in the manuscript

## 4 DISCUSSION

The `MetaRVM` model offers an important advancement in epidemiological modeling by providing a flexible framework for simulating infectious disease spread across stratified subpopulations. This model effectively balances the computational simplicity of compartmental models with the detailed granularity of agent-based models, making it suitable for quick-turnaround public health decision support. Our collaboration with CDPH has been instrumental in ensuring that `MetaRVM` is responsive to real-world needs and integrates into existing public health workflows. Through direct engagement with CDPH, we have tailored `MetaRVM` to address specific challenges faced by public health officials in Chicago and similar densely populated regions amid varying contact patterns enforced by the diverse demographic. This partnership has facilitated the co-creation of a model that is operationally viable for routine use. The model’s ability to simulate disease dynamics across different age groups and geographic areas can be valuable for tracking influenza and other respiratory disease dynamics and informing intervention strategies.

Despite its strengths, the `MetaRVM` model relies heavily on the quality of input data, including the mixing matrices and detailed vaccination. Inaccuracies and uncertainties in these data sources may impact the quality of the model’s projections. Additionally, while `MetaRVM` captures key aspects of disease progression, it does not simulate individual-level interactions, which may limit its ability to model highly localized transmission dynamics. Continued collaboration with CDPH and other stakeholders will be crucial to refining the model and ensuring its relevance in diverse epidemiological contexts. In summary, the `MetaRVM` model represents a collaborative endeavor that enhances public health analytics in Chicago, offering a practical tool for understanding and managing infectious disease outbreaks.

## Acknowledgments

This material is based upon work supported by the Chicago Department of Public Health. This research was completed with resources provided by the Laboratory Computing Resource Center at Argonne National Laboratory (Bebop cluster).

## Notes

### Competing Interest Statement

The authors have declared no competing interest.

